# Navigating the chaos of psychedelic fMRI brain-entropy via multi-metric evaluations of acute psilocybin effects

**DOI:** 10.1101/2023.07.03.23292164

**Authors:** Drummond E-Wen McCulloch, Anders Stevnhoved Olsen, Brice Ozenne, Kristian Larsen, Dea Siggaard Stenbæk, Sophia Armand, Martin Korsbak Madsen, Gitte Moos Knudsen, Patrick MacDonald Fisher

**Affiliations:** Neurobiology Research Unit, Copenhagen University Hospital, Rigshospitalet, Copenhagen, Denmark; Faculty of Health and Medical Sciences, University of Copenhagen, Denmark; Department of Applied Mathematics and Computer Science, Technical University of Denmark, Kgs. Lyngby, Denmark; Section of Biostatistics, Department of Public Health, University of Copenhagen, Copenhagen, Denmark; Department of Psychology, University of Copenhagen, Denmark; Department of Psychiatry Svendborg, Svendborg, Denmark; Department of Clinical Medicine, University of Copenhagen, Copenhagen, Denmark; Department of Drug Design and Pharmacology, University of Copenhagen, Copenhagen, Denmark

## Abstract

A prominent theory of psychedelics is that they increase brain entropy. Thirteen studies have evaluated psychedelic effects on fMRI brain entropy; no findings have been replicated. Here we evaluated these metrics in an independent 28-participant healthy cohort with 121 pre- and post-psilocybin fMRI scans. We assessed relations between brain entropy and objective and subjective psychedelic drug effects using linear mixed-effects models. All metrics were evaluated using two parcellation strategies and 7 denoising pipelines. We observed consistent significant positive associations for Shannon entropy of the spatial eigendistribution of the time by voxel matrix, path-length, instantaneous correlations, brain-state switching, and sample entropy at short time-scales. We consistently did not observe significant effects for 8 of 14 entropy metrics and observe inconsistent positive effects for Lempel-Ziv complexity of the BOLD signal. Brain entropy quantifications showed limited inter-measure correlations. Our observations support a nuanced acute psychedelic effect on brain entropy, empirically demonstrating that these metrics do not reflect a singular construct.

## Introduction

Psychedelic drugs induce profound altered states of consciousness including affective, sensory and cognitive effects mediated by activation of downstream pathways initiated by agonism at the brain serotonin 2A receptor ^1–4^. In combination with psychological support, clinical studies up to phase 2b indicate promising clinical efficacy of psychedelics in the treatment of affective and behavioural neuropsychiatric disorders that may be associated with their acute effects ^5–10,10–12^. Similarly, psychedelics induce acute and lasting effects on behaviour and personality in healthy participants ^13–15^. In parallel to evaluating clinical treatment effects, human brain functional magnetic resonance imaging (fMRI) has begun to shed light on neural pathways affected by psychedelics ^16^.

Several prominent theories of critically relevant psychedelic effects on brain function have been advanced^17^. A prominent theory is the “Entropic Brain Hypothesis” (EBH), which posits that the ‘richness’ of the phenomenology of the acute psychedelic state reflects brain-wide increases in entropy of functional brain signals ^18,19^. The concept of "entropy" serves as a quantification of the degree of information or complexity contained in a system; it is typically expressed in bits, without physical dimensions. Entropy has been conceptualised as a metric of information content and is largely defined in terms of a probability distribution; the metric is commonly referred to by the eponymous name “Shannon entropy” *H*(*X*) = − ∑_*x*∈*X*_ *p*(*x*) ⋅ *log*_2_*p*(*x*) where H(X) refers to the Shannon entropy of probability mass function X containing bins (x) with probability (p) ^20^. Other entropy metrics have been defined, e.g., Lempel-Ziv complexity and sample entropy ^21–24^, and adapted to characterise information contained in various data representations, e.g., complex networks or graph structures ^25^.

To date, 13 studies have evaluated either acute or lasting psychedelic effects on the information-entropy of brain activity or connectivity using blood oxygen level dependent (BOLD) fMRI data (Figure 1), one of which evaluates two metrics, thus 14 entropy metrics have been previously evaluated in this field. Four papers analysed data from a study evaluating 2mg intravenous psilocybin administration in up to 15 healthy participants ^19,26–28^. Three papers reported effects from a study evaluating 75µg intravenous LSD administration in 15 healthy participants ^29,30^, one of which also evaluated the effect of 25 mg oral psilocybin in 6 healthy participants ^31^. Two papers evaluated data from both intravenous datasets ^32,33^. Three papers reported effects from a study evaluating oral ayahuasca administration, containing 96-160mg DMT and 25.2-42mg harmine (a monoamine oxidase inhibitor that facilitated the oral bioavailability of DMT) in 9 healthy participants ^34–36^. Finally, one paper reported effects from a study evaluating lasting effects of 25mg/70kg bodyweight orally administered psilocybin in 11 healthy participants ^37^. Notably, and highlighted in a recent review ^16^, each of these reports quantified a distinctly different metric of brain entropy. Here we group these metrics into three categories: 1) “static connectivity”, 2) “dynamic connectivity”, i.e., the time-varying relation between two or more time-series, 3) “dynamic activity”, i.e., the entropy of regional time-series (Figure 1). Ten of these metrics are based on the Shannon entropy of distributions, three are Lempel-Ziv complexity metrics of a time-series and one is the sample entropy of a time-series. Taken together, although there is a clear interest in evaluating psychedelic effects on brain entropy, prominent limitations include that none of these measures have been evaluated in an independent cohort, the set of effects have been evaluated in only a few datasets, some with atypical modes of drug administration, and the inter-correlation between these metrics has not been considered. Previous studies compared pre-drug or placebo and a single post-drug scan. For the most part, these previous studies report increased brain entropy with only a few exceptions, e.g., two studies report non-significant psilocybin effects and one study reports no lasting changes (Figure 1, Table 1). See Supplementary Table S1 for more details about the previous studies. Previous fMRI studies have reported reduced brain entropy in other states of "reduced consciousness", including NREM sleep ^38^, minimal-consciousness ^39^, and anaesthesia ^40^, whereas other studies have shown increased brain entropy following caffeine ^41^ and Salvinorin A intake ^42^. In the current study, we sought to evaluate the acute effects of 0.2-0.3 mg/kg psilocybin administration on these 14 brain entropy metrics in an independent dataset of 28 healthy individuals. Based on the EBH, we hypothesised that brain entropy metrics would be increased following psilocybin administration. Participants completed a 5 or 10-min resting-state fMRI scan a single time before, and multiple times following psilocybin administration (121 total scan sessions). All scans for each participant were performed on one of two scanners. Each scan was accompanied by a self-report measure of subjective drug intensity (SDI) and a blood sample to quantify plasma psilocin level (PPL), from which brain serotonin 2A (5-HT2A) receptor occupancy (Occ_2A_), was estimated based on its relation to PPL established in a previous study from our lab ^2^. We evaluated the relations between each entropy metric and SDI, PPL and Occ_2A_ using linear mixed-effect models with a subject-specific random intercept and correction for age, sex, scanner, and motion (see Methods Supplementary Material for more details). We report uncorrected permutation p-values (p_perm_) for each metric producing a single whole-brain value. For entropy metrics producing regional values, we report family-wise error rate corrected permutation p-values (p_FWER_) with correction across all regions within each metric using maxT correction ^43^. We report as significant those metrics which were significantly (i.e., p < 0.05) associated with SDI, PPL and Occ_2A_, collectively referred to as “PsiFx”. Standardised effect sizes are reported (Pearson’s rho). This evaluation was repeated across two parcellation strategies and seven pre-processing pipelines to explore the robustness of effects to pre-processing decisions. Finally, we explored the inter-correlation between brain entropy metrics to characterise their associations.

**Figure 1:**
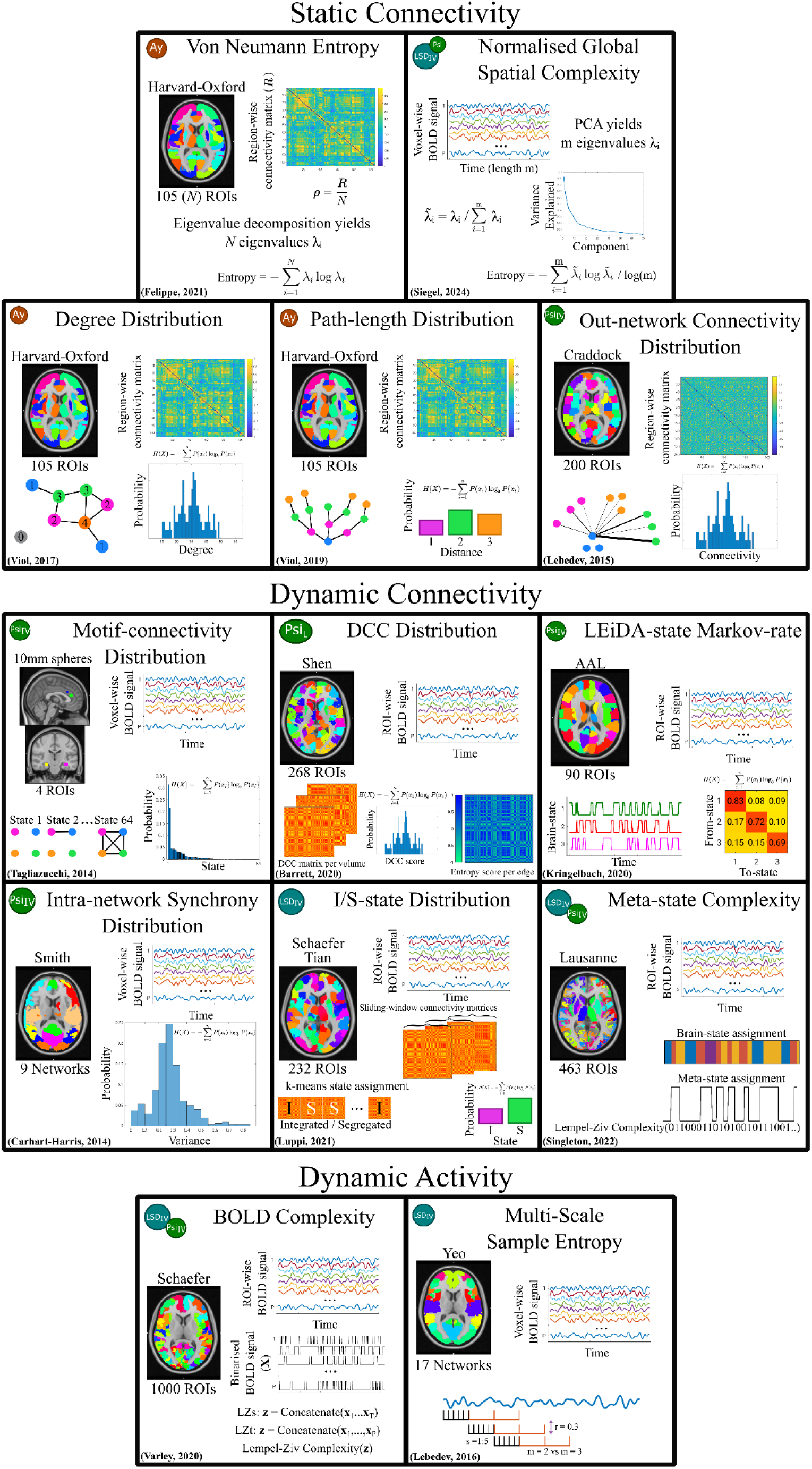
Overview of previous psychedelic fMRI entropy quantifications. Methods are grouped according to the type of brain entropy metric. Symbols in the top left of each box represent the dataset that the original publication analysed with icon area scaled by sample size. LSD_IV_ (blue) refers to data collected in relation to a 75 µg intravenous LSD administration (n <= 15), PsiIV (green) refers to data collected in relation to a 2 mg intravenous psilocybin administration (n <= 15), Psi (green) refers to data collected in relation to a 25 mg oral psilocybin administration (n = 6), Ay (orange) refers to data collected in relation to an oral ayahuasca administration (n = 9), and Psi_L_ (green oval) refers to data collected one-week before, one-week after, and one-month after a 25 mg/70 kg oral psilocybin administration (n = 11). Brain images show an axial slice illustrating the parcellation/atlas used. Images in the top right describe the input into the entropy function as one of "region-wise connectivity matrix", "ROI-wise BOLD signal" and "voxel-wise BOLD signal". Illustrations in the lower part of each box graphically represent simplified analysis steps for each method. The original publication for each metric is denoted in the bottom left corner of each box. See Supplementary Table 1 for more details.

**Table 1:** Summary of entropy quantification methods, original findings, and findings reported within this manuscript. For the “Original findings” and “Our findings” columns, grey cells describe no association of effect, light grey refers to no reporting of acute effects on brain entropy, yellow describes marginal effects, and green or blue statistically significant effects. For a more in-depth evaluation of our findings please see supplementary tables S1, S2, S8 and S9.

| Entropy | Original dataset | Original findings | Our findings (original atlases & generic denoising) | Our findings (across 6 denoising strategies) |
| --- | --- | --- | --- | --- |
| **Static Connectivity** |  |  |  |  |
| Out-network connectivity distribution entropy | IV Psi | Region-specific effects | Not associated with PsIFx | Positive association in 1/181 regions in 1/6 pipelines (moderate)<br>Negative association in 1/181 regions in 1/6 pipelines (weak) |
| Degree distribution entropy | Oral Aya | Increased | Not associated with PsIFx | Positive associations at sparse specific degrees in 3/6 pipelines (weak to moderate) |
| Geodesic distribution entropy | Oral Aya | Increased | Weak-moderate association with PsIFx† | Positive associations in 5/6 pipelines (weak to moderate)<br>Negative association in degrees 7-9 following GSR (weak) |
| Von Neumann entropy | Oral Aya | Numerically increased | Not associated with PsIFx | Positive associations with some PsIFx in 3/6 pipelines (weak to moderate) |
| Normalised Global Spatial Complexity | Oral Psi & IV LSD | Increased | Moderate association with PsIFx | Positive associations with all PsIFx in all 6/6 pipelines (moderate) |
| **Dynamic Connectivity** |  |  |  |  |
| Intra-network synchrony distribution entropy | IV Psi | Increased (some networks) | Not associated with PsIFx | Negative association in 2/9 networks in 1/6 pipelines (moderate) |
| Motif-connectivity distribution entropy | IV Psi | Increased | Not associated with PsIFx | Positive association in 1 or 2/75 windows in 3/6 pipelines (weak)<br>Negative association in 5/75 windows in 1/6 pipelines (weak to moderate) |
| Meta-state complexity | IV Psi & IV LSD | No change | Weak association with SDI and Occ but not PPL | Positive associations with some PsIFx in 5/6 pipelines (weak) |
| I/S state distribution entropy | IV LSD | No change | Not associated with PsIFx | Never associated with PsIFx |
| LEIDA state Markov Rate entropy | IV LSD | Not reported | Not associated with PsIFx | Positive association with SDI and Occ only in 1/6 pipelines (weak) |
| Edge-wise DCC distribution entropy | Oral Psi* | No persisting change | Moderate-strong association with all PsIFx all networks except within motor cortex | Positive associations with all PsIFx in 4/6 pipelines (moderate to strong)<br>Negative associations in 8/36 edges when lowpass filter removed (weak)<br>No association following narrowbandpass. |
| **Dynamic Activity** |  |  |  |  |
| Sample entropy (4-6 seconds) | IV LSD | Increased at scales 1, 2 and 3 | Weak-moderate association with all PsIFx in several networks | Positive associations with all PsIFx in 2/6 pipelines (moderate to strong)<br>Negative associations when lowpass filter removed (weak to moderate) |
| Multi-scale sample entropy (20-30 seconds) | IV LSD | Decreased at scale 5 | Weak-Moderate negative association with PsIFx in several networks scale 5 | Negative association in 1/6 pipelines (weak to moderate) |
| BOLD complexity (spatial) | IV Psi & IV LSD | Increased (LSD)<br>No change (psilocybin) | Not associated with PsIFx | Negative association with Occ only when lowpass filter removed |
| BOLD complexity (temporal) | IV Psi & IV LSD | Not reported | Weak association with SDI and Occ but not PPL | Positive associations with all PsIFx in 2/6 pipelines (moderate) |

\*Investigates persisting effects at 1 week and 1 month post-psilocybin administration
† at thresholds producing a mean degree of 22-38

## Results

Participants showed substantial SDI and PPL following drug administration as anticipated (Supplementary Figure S1). See Figure 1 and Table 1 for a summary of entropy metrics, our findings, and previous findings. Primary analyses used the spatial parcellation reported in the original publication (with cerebellum removed) and a generic denoising pipeline (12 motion parameters, aCompCor, interpolation (scrubbing) of artefactual volumes, and bandpass filtering in the range 0.008-0.09Hz). To explore moderating effects of denoising pipelines on brain entropy metrics, we considered six variations 1) including global-signal regression (GSR), 2) removal of the low-pass filter (0.09 Hz), 3) applying a narrow bandpass filter (0.03-0.07 Hz), 4) regressing 24 motion parameters, 5) omitting scrubbing, and 6) a stricter scrubbing threshold. For a summary of results across denoising pipelines see Supplementary Table S8 and for a full breakdown across all pipelines see Supplementary Table S9.

Here we show a significant and consistent relation between objective (i.e., PPL) and subjective (i.e., SDI) measures of psilocybin effects and five of the 14 entropy metrics evaluated, inconsistent but positive effects for one, and consistently null effects for eight metrics. Those demonstrating a consistent relation with psilocybin effects across denoising pipelines were distributed across entropy classes: two features of static connectivity, two of dynamic connectivity and one of dynamic activity. No single entropy metric stands out as the clearest candidate for future research. Our observation that the metrics are generally weakly correlated with each other emphasises that they represent distinct phenomena, i.e., “brain entropy” is not a singular construct.

### Entropy of Static Connectivity

#### Out-network Connectivity Distribution Entropy

Shannon entropy of regional distribution of out-network correlation coefficients was not significantly associated with psilocybin effects (PsiFx) in any of the 181 non-cerebellar brain regions after controlling for multiple comparisons (p_FWER_ > 0.07 for all regions for at least one of PsiFx, Supplementary Table S2). Across additional denoising pipelines, only one region had a moderate significant positive relation and one had a weak significant negative relation, each for only a single pipeline (Supplementary Tables S8, S9).

#### Degree Distribution Entropy

Shannon entropy of global degree distribution at a correlation coefficient threshold corresponding to a mean degree of 27 was not associated with any of PsiFx (p_perm_ > 0.18, Figure 2A). We also did not observe significant effects for thresholds producing a mean degree between 1 and 48 (Supplementary Table S3). Across additional denoising pipelines, there were sparse weak-to-moderate significant positive associations in three of six pipelines, with little overlap in which degrees were significant (Supplementary Tables S8, S9).

**Figure 2:**
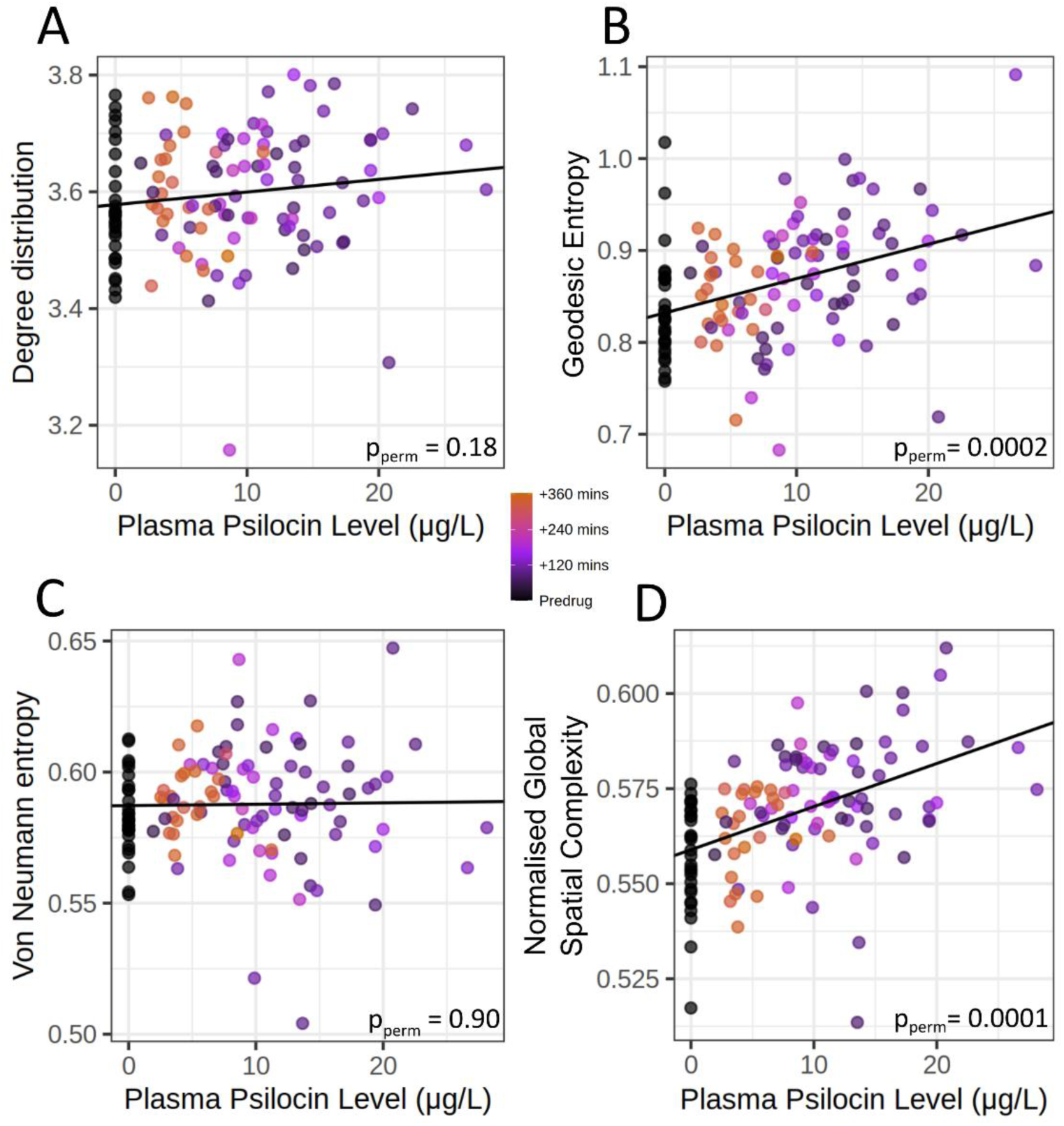
Scatter plots describing the relation between static connectivity entropy and PPL. Y-axis values are partial residuals i.e., entropy values adjusted for age, sex, MR scanner and motion. Degree distribution and Geodesic distribution entropy statistics are computed using a correlation coefficient threshold corresponding to a mean-degree of 27. Each scatter plot includes repeated measures (n = 121 scans) for each subject (n = 28 subjects).

#### Geodesic Distribution Entropy

The entropy of path-length (geodesic) distribution was significantly positively associated with PsiFx at the a priori described threshold producing mean degree 27 (p_perm_ < 0.04, Figure 2B). The associations were weak to moderate (Pearson’s rho = 0.39, 0.27, and 0.23 for PPL, Occ_2A_ and SDI, respectively). Significant weak to moderate positive associations with PsiFx were also observed across thresholds producing mean degrees from 22 to 38 (Supplementary Table S3). This weak-to-moderate positive effect was consistent across five of six additional denoising pipelines, the exception being GSR (Supplementary Tables S8, S9).

#### Von Neumann Entropy

The Von Neumann entropy of correlation matrices was not significantly associated with PsiFx in the primary analysis (p_perm_ > 0.35, Figure 2C and Supplementary Table S3). However, weak-to-moderate significant positive effects were observed in three of six denoising pipelines for some PsiFx, with the strongest relation following GSR (Pearson’s rho = 0.43, 0.52 and 0.36 for PPL, Occ_2A_ and SDI, respectively) (Supplementary Tables S8, S9).

#### Normalized Global Spatial Complexity

Normalized Global Spatial Complexity (NGSC) was significantly moderately positively associated with PsiFx (p_perm_ < 0.0003, Figure 2D and Supplementary Table S3; Pearson’s rho = 0.47, 0.44, and 0.52 for PPL, Occ_2A_ and SDI, respectively). This relation was consistent across all denoising pipelines (Supplementary Tables S8, S9).

### Entropy of Dynamic Connectivity

#### Intra-network Synchrony Distribution Entropy

Intra-network synchrony distribution entropy was not significantly associated with PsiFx in any of nine networks in the primary analysis (All p_FWER_ > 0.98, Supplementary Table S2, Supplementary Figure S2). Following the removal of the lowpass filter, the visual and somatomotor networks showed a significant moderate negative relation with all PsiFx (Supplementary Tables S8, S9).

#### Motif-connectivity Distribution Entropy

The four-ROI motif-connectivity state distribution was not significantly associated with PsiFx at any window length from 15 to 150s, except a single weak association at window-length 100s (p_perm_ < 0.05, Pearson’s rho 0.30, 0.25, and 0.24 for PPL, Occ_2A_, and SDI, respectively) surrounded by non-significant findings (Supplementary Figure S3 and Supplementary Table S3, S9). Across only four denoising pipelines, only few individual window lengths were significantly associated with PsiFx, some of which were positive and some negative (Supplementary Tables S8, S9).

#### LEiDA-state Markov-rate Entropy

LEiDA-state Markov-rate was not significantly associated with PsiFx (p_perm_ > 0.7 for all PsiFx, Figure 3A, Supplementary Table S3, S9). A weak positive relation with SDI and Occ_2A_ was observed only following GSR (Pearson’s rho = 0.26 and 0.21, respectively) (Supplementary Tables S8, S9).

**Figure 3:**
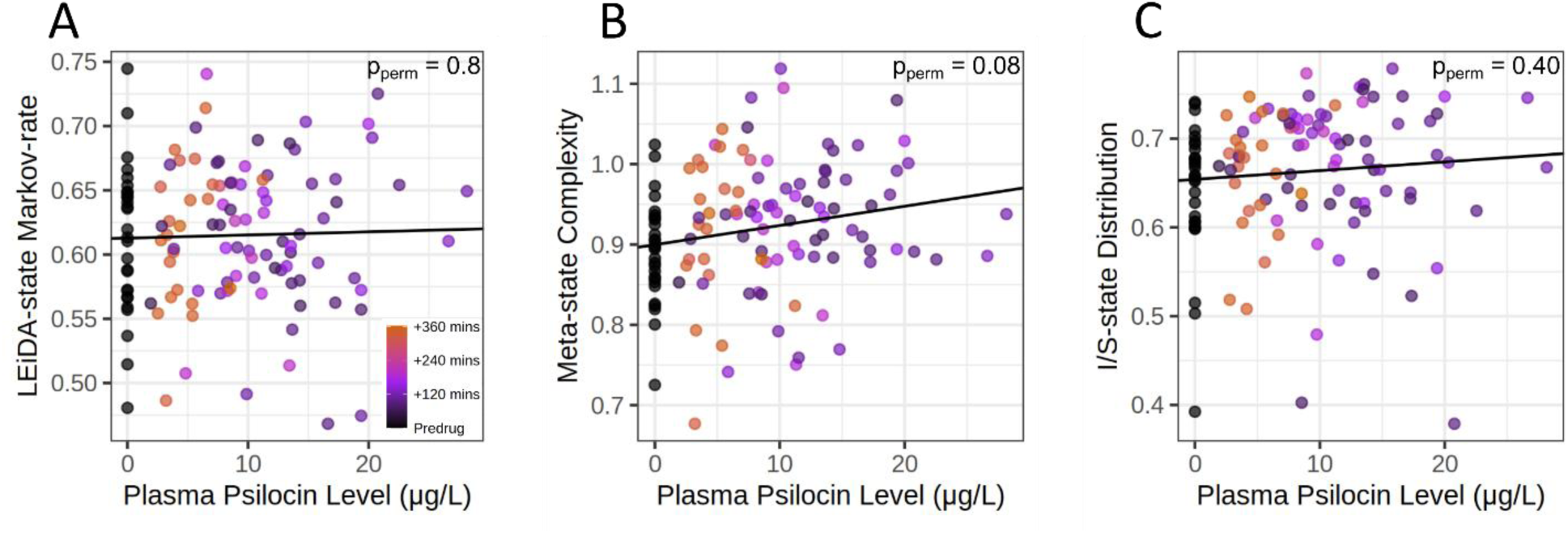
Scatter plots and linear models describing the relation between whole-brain dynamic connectivity entropy measures and plasma psilocin levels. Y-axis values are partial residuals i.e., entropy values adjusted for age, sex, MR scanner and motion. Panel B (Meta-state complexity) shows a non-significant association with PPL but this entropy metric does show a significant linear relation with Occ_2A_ and SDI. Each scatter plot includes repeated measures (n = 121 scans) for each subject (n = 28 subjects).

#### Meta-state Complexity

Meta-state complexity was positively associated with Occ_2A_ (p_perm_ = 0.03) and SDI (p_perm_ = 0.003), but not PPL (p_perm_ = 0.076, Figure 3B). Associations were weak (Pearson’s rho = 0.22, 0.33, and 0.20 for Occ_2A_, SDI, and PPL, respectively; Supplementary Table S3, S9). The weak positive associations remained significant across five of the six additional denoising pipelines (Supplementary Tables S8, S9).

#### Integration/Segregation-state Distribution Entropy

Integration sub-state entropy was not significantly associated with PsiFx in either the primary or additional pipelines (p_perm_ > 0.06 for all PsiFx, Figure 3C, Supplementary Table S3, S8, S9).

#### Dynamic Conditional Correlation Distribution Entropy

Dynamic conditional correlation (DCC) entropy was significantly positively associated with PsiFx in 35 of 36 network-network connections (18/36 p_FWER_ < 0.0001, i.e., observed data superseded all permutations, 29/36 p_FWER_ < 0.001, 35/36 p_FWER_ < 0.05; Figure 4A; Supplementary Table S2). Associations were moderate to strong (Pearson’s rho range: 0.35 to 0.78, Supplementary Table S2). The one association with at least one non-significant relation was for edges within the motor cortex. The positive associations replicated in four of six denoising pipelines. DCC entropy effects were particularly sensitive to temporal filtering: employing a narrow bandpass filter removed the associations and removing the low-pass filter changed to negative the association for 8 of 36 network-network connections. (Supplementary Tables S8, S9).

**Figure 4:**
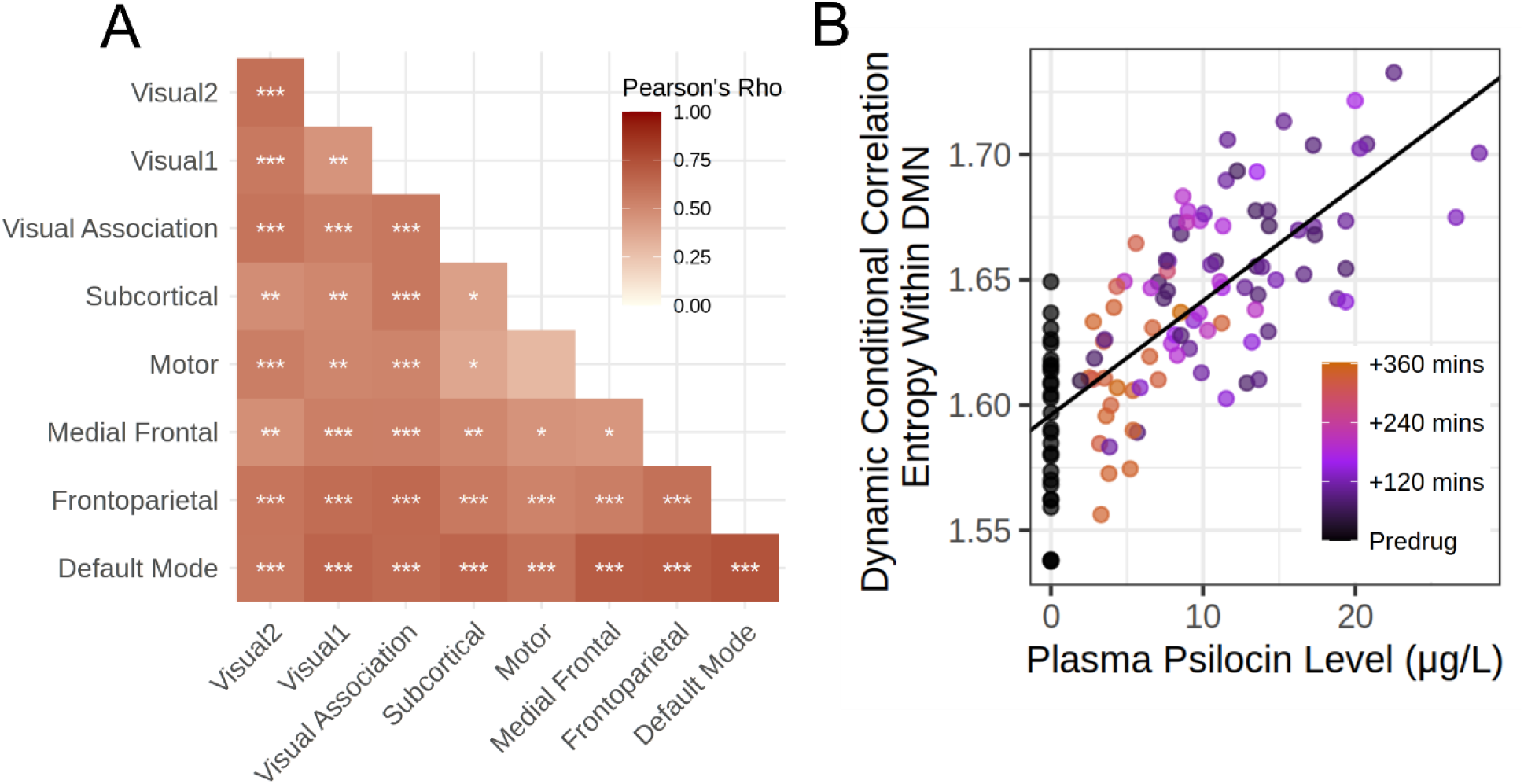
**A:** Heatmap of the Pearson’s correlation and p-values for the association between Dynamic Conditional Correlation entropy and plasma psilocin level for each within and between network entropy estimate. *** represents p_FWER_ < 0.0001, ** p_FWER_ < 0.001, and * p_FWER_ < 0.05 for associations with PPL. **B:** A scatter plot of the network edge with the strongest association between DCC entropy and PPL (Pearson’s Rho = 0.74). Y-axis values are partial residuals i.e., entropy values adjusted for age, sex, MR scanner and motion. The scatter plot includes repeated measures (n = 121 scans) for each subject (n = 28 subjects).

### Entropy of Dynamic Activity

#### Multi-Scale Sample Entropy

At scale 1, (i.e., no time-series compression), sample entropy was significantly positively associated with PsiFx (p_FWER_ < 0.05) in 7 of 17 networks (i.e., Central Visual, Dorsal Attention A, Control A, B and C, Default-Mode A and C). This finding replicated only in two of six additional denoising pipelines. When the low-pass filter was removed, negative associations were observed (Supplementary Tables S8, S9). At scales 2, 3, and 4, no associations were significantly associated with PsiFx (p_FWER_ > 0.05). At scale 5, sample entropy was significantly negatively associated with PsiFx in 14 of 17 networks; Control A and C and Default-Mode A (p_FWER_ < 0.001), Somatomotor A, Dorsal Attention A and B, Salience-Ventral-Attention B, Limbic B, Control B, Temporal-Parietal, Default-Mode B and C (p_FWER_ < 0.05, Figure 5, Supplementary Table S2). Scale 1 associations were weak to moderate (Pearson’s rho range: 0.26 to 0.47), as were Scale 5 associations (Pearson’s rho range: -0.27 to -0.49). The scale 5 finding only replicated for one additional denoising pipeline, i.e., removal of the lowpass filter (Supplementary Tables S8, S9).

**Figure 5:**
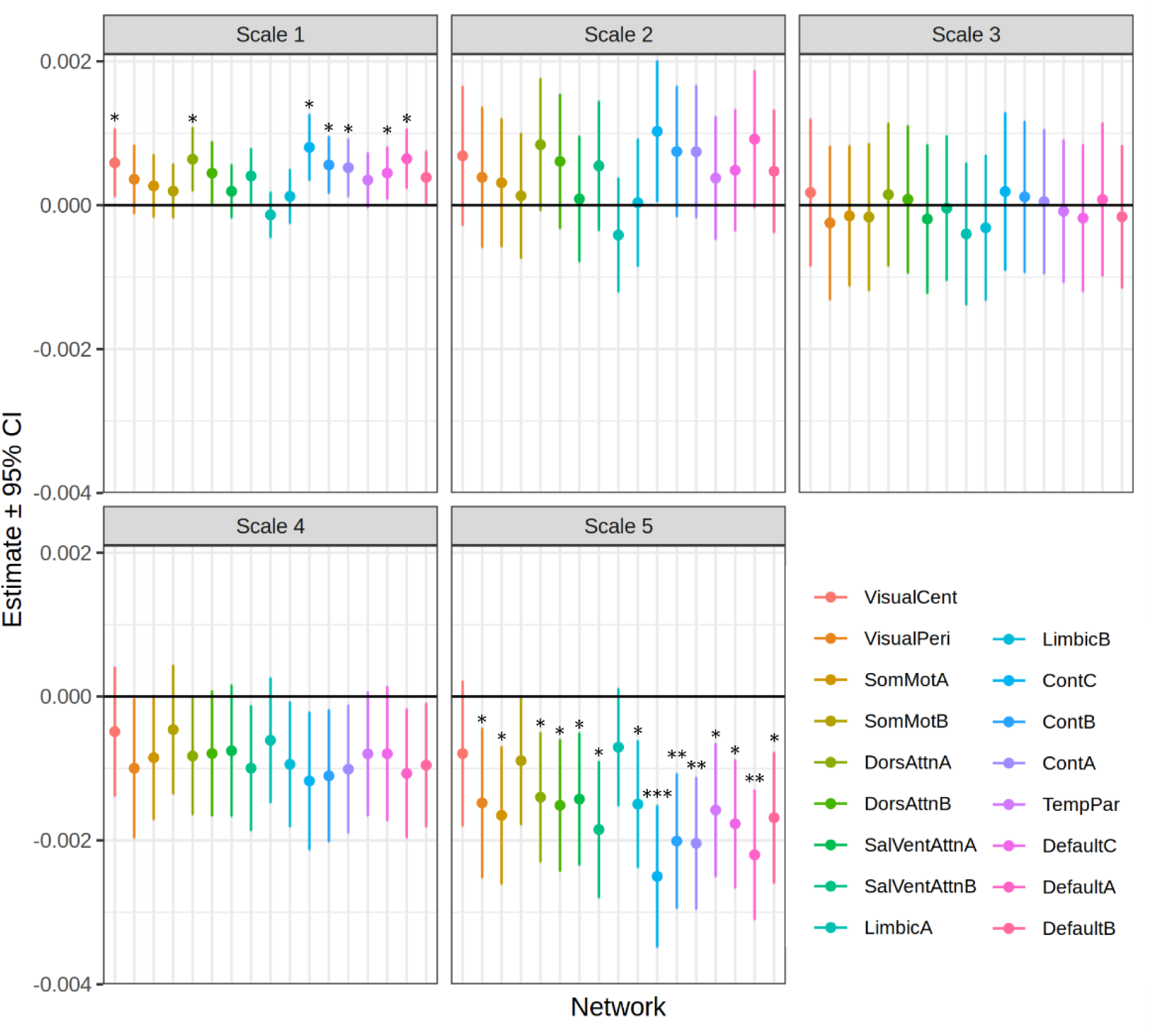
Forest plots representing the estimate of the association between sample entropy and PPL in each network of the Yeo-17 network parcellation and at each of scales 1 to 5. Colours represent networks and error bars represent the 95% confidence interval. *** p_FWER_ < 0.0001, ** p_FWER_ < 0.001 and * p_FWER_ < 0.05 for associations with PPL.

#### Spatial and Temporal Dynamic BOLD Complexity

Temporal BOLD complexity (LZct) was not associated with PPL (p_perm_ = 0.14, Figure 6B) but was associated with Occ_2A_ (p_perm_ = 0.03) and SDI (p_perm_ = 0.009). Associations were weak (Pearson’s rho = 0.23, 0.30 and 0.17 for Occ_2A_, SDI, and PPL respectively). Spatial BOLD complexity (LZcs) was not significantly associated with PsiFx (p_perm_ > 0.6, Figure 6A, Supplementary Table S3. LZct was significantly moderately associated with all PsiFx in two of six denoising pipelines, i.e., no scrubbing and strict scrubbing. LZcs was not associated with PsiFx for any pipeline (Supplementary Tables S8, S9).

**Figure 6:**
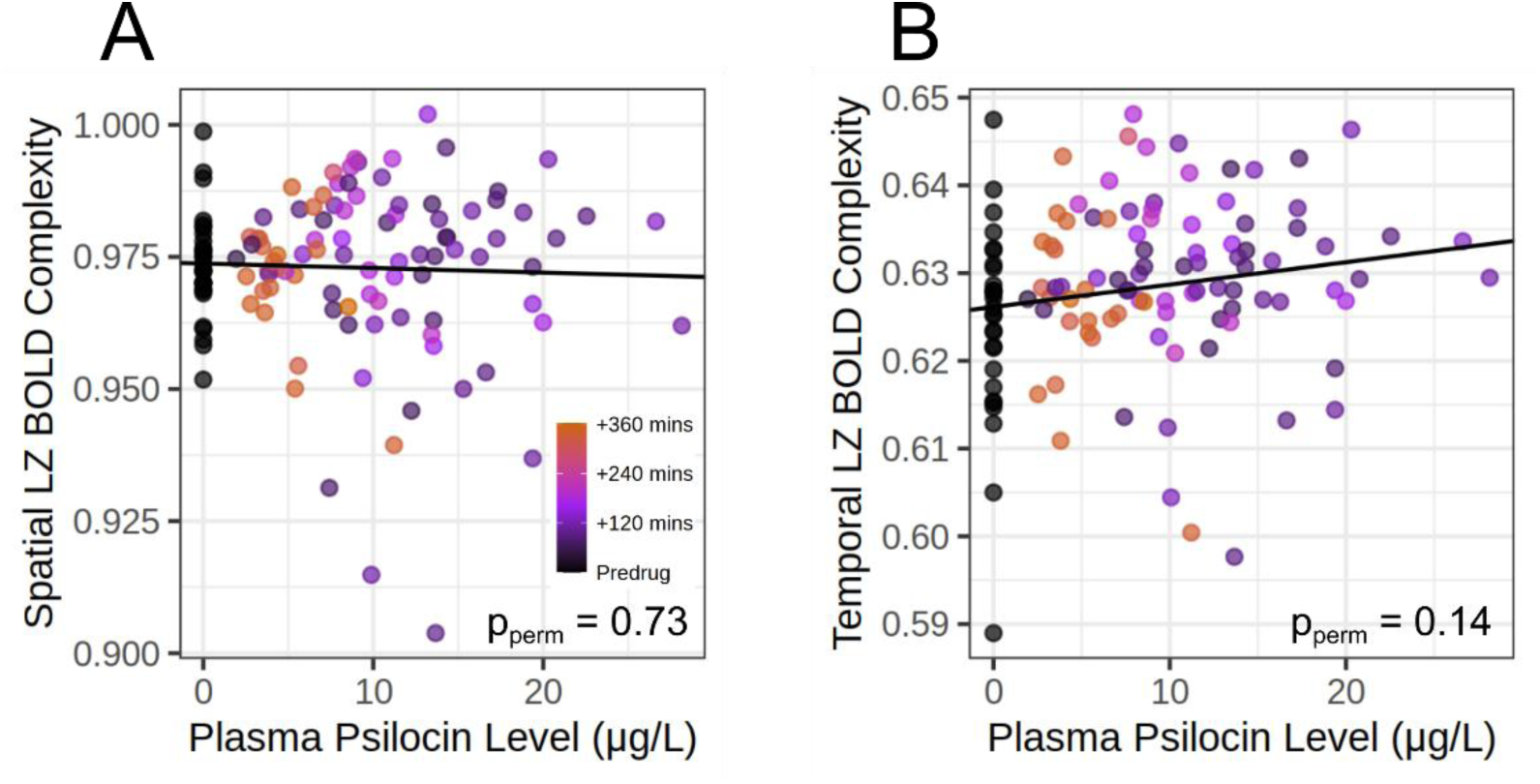
Scatter plots and linear models describing the relation between spatial (A) and temporal (B) Lempel-Ziv entropy of dynamic BOLD activity and PPL. Y-axis values are partial residuals i.e., entropy values adjusted for age, sex, MR scanner and motion. Temporal BOLD complexity (LZct) was significantly, but weakly associated with both Occ_2A_ and SDI despite not being significantly associated with PPL. Each scatter plot includes repeated measures (n = 121 scans) for each subject (n = 28 subjects).

### Effect of Parcellation

Main analyses were performed using the same atlas as the original publication. To evaluate PsiFx on brain entropy metrics using a common parcellation, all analyses using the original denoising pipeline were re-run using an atlas combining the Schaefer-100 (7 Yeo networks) and Tian-16 subcortical atlases^44,45^, see Supplementary Table S5 for detailed results. Geodesic entropy showed a weak positive association with all PsiFx at mean degrees 31 to 38. Meta-state complexity was weakly positively associated with Occ, but not PPL nor SDI. LEiDA-state Markov-rate was weakly negatively associated with all PsiFx and DCC distribution was weak-to-strongly associated with all PsiFx across most network edges. Sample Entropy was weak-to-moderately associated with PsiFx at scale 1 but no significant associations were observed at longer scales, although the trend of increased entropy at short scales and decreased entropy at long-scales was maintained. No parcellation was applied for NGSC (voxel-wise) or Motif-connectivity Distribution Entropy (uses predefined seeds). All other metrics were not significantly associated with PsiFx.

### Moderating Effect of Scanner

To evaluate scanner effects on observed associations from the main analysis pipeline, we fit linear mixed models estimating the moderating effect of scanner, see Supplementary Table S6 for detailed Results. We observed a significant moderating effect of scanner on the relation between PPL and entropy for geodesic entropy over the range of thresholds at which we observe significant associations with PsiFx. The nature of this interaction was that for scanner A the effect was closer to zero than for scanner B. We also show a significant moderating effect of scanner for Sample Entropy scale 5 across many ROIs that are significantly associated with PsiFx. However, the observed effect is the same direction for scanner A and B, only numerically stronger for scanner A. We observed a significant moderating effect of scanner for Von Neumann entropy such that the associations with PsiFx were opposite for the two scanners. We did not observe a significant moderating effect of scanner for DCC entropy, nor NGSC, supporting the robustness of these metrics. For entropy metrics that were not associated with PsiFx in the main model, we observed moderating effects of scanner for I/S-state distribution, some window-lengths of motif-connectivity distribution, some ROIs of out-network connectivity, degree distribution, and some sample entropy scale 4 ROIs.

### Correlation Between Whole-brain Entropy Quantifications

We estimated the correlation between whole-brain entropy metrics to explore their association with one another across all included scans from the main analysis pipeline. Some metrics were positively and some negatively correlated with other metrics and some pairs were not correlated with one another (Figure 7). After correction for multiple comparisons, six entropy quantification pairs were positively related (LZcs & LZct, LZcs & Von Neumann, geodesic entropy & degree-distribution, LEiDA state & Von Neumann, motif connectivity distribution-window100 & Von Neumann, NGSC & Von Neumann) and four were negatively related (geodesic & LEiDA state, geodesic & Von Neumann, degree-distribution & LEiDA state, degree-distribution & Von Neumann, NGSC & geodesic). See Supplementary Table S4 for pairwise correlation coefficients and p-values.

**Figure 7:**
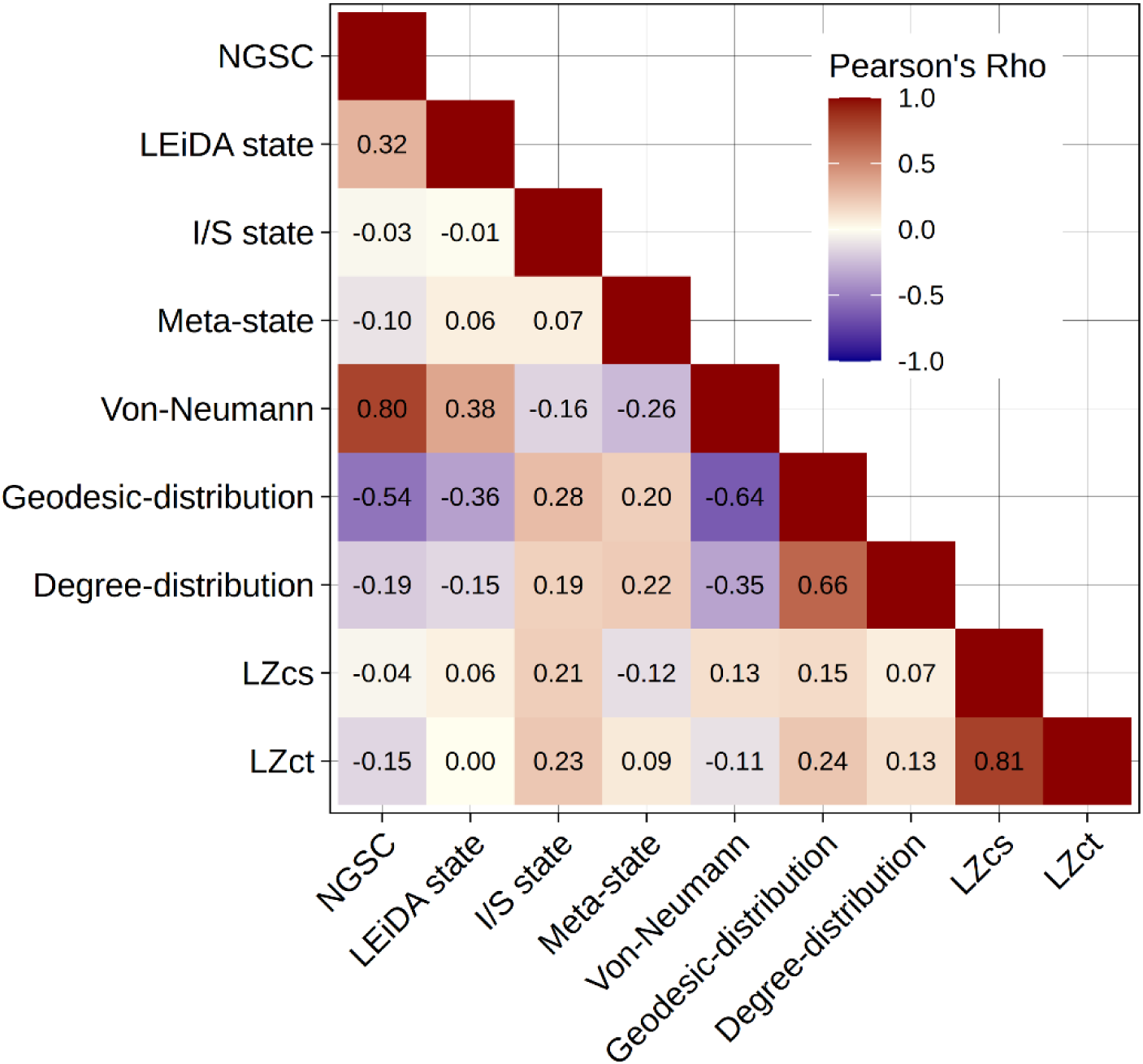
Heatmap showing the correlation coefficients between whole-brain entropy metrics. Colours represent the Pearson’s correlation coefficient. NGSC, Normalised Global Spatial Complexity; I/S state, Integration/Segregation state distribution; MCD, Motif connectivity distribution (with either 100 or 15 second windows); LZcs, Spatial Lempel-Ziv complexity. LZct, Temporal Lempel-Ziv complexity.

## Discussion

### Overview

Recent studies have reported acute psychedelic effects on functional brain entropy, but to date none of these metrics have been independently reevaluated. In this study we evaluated 14 previously reported entropy metrics in a novel sample of 28 healthy participants scanned with BOLD fMRI before and several times after psilocybin with concomitant measurements of subjective drug intensity and plasma psilocin level. We observed statistically significant psilocybin effects that echoed previous reports for three brain entropy metrics: geodesic entropy, wherein we replicate increased entropy at previously reported thresholds; sample entropy, wherein we replicate a previously observed increase in entropy at short scales and decrease at long scales; and NGSC where we replicate previously observed increases. We observed a strong positive relation between PsiFx and brain entropy measured by Dynamic Conditional Correlation analyses that has not been previously reported. Meta-state complexity showed significant relations with PsiFx across most preprocessing pipelines, and temporal Lempel-Ziv complexity showed evidence for associations with PsiFx across some pipelines. For 8 of 14 brain entropy metrics previously reported, we did not observe a significant association with psilocybin measures, and we see limited correlation between entropy metrics. These mixed findings underscore the importance of corroborating outcomes in independent datasets. Although we observe some evidence supporting the entropic brain hypothesis, these variable findings underscore the broadness of this theory and the need to more clearly establish which brain entropy metrics of functional brain imaging signals are acutely affected by psychedelics.

### Normalised Global Spatial Complexity

We found that NGSC, the entropy of (spatial) eigenvalues of z-scored voxel-wise data, was significantly positively associated with PsiFx across denoising pipelines. This is consistent with the original report from Siegel et. al.,^31^ who report increases following oral psilocybin and intravenous LSD using the publicly available dataset ^46^. This indicates that psilocybin increases the uniformity of variance distribution across spatial principal components. Not noted previously, NGSC and Von Neumann entropy are nearly mathematically equivalent, Von Neumann entropy uses a parcellated, region-wise instead of voxel-wise, correlation matrix. Unsurprisingly, they are strongly positively correlated in our sample (Pearson’s rho = 0.81). Despite this, Von Neumann entropy was not associated with PsiFx, except after GSR, implying that GSR affects parcellation more than voxel-wise connectivity matrices. Although reducing spatial dimensionality may be necessary for comparison of brain regions across participants, it may inadvertently average away complexity observable at the voxel level. Future studies exploring these trade-offs could better highlight when one method or the other is preferred. Our results provide evidence for NGSC as a sensitive biomarker of psychedelic drug effects, robust to denoising strategy. However, we are not aware of any fMRI studies evaluating the effect of other psychoactive drugs on NGSC so cannot infer specificity.

### Geodesic Entropy

We report a significant positive association between the Shannon entropy of the distribution of path lengths across the whole brain as previously reported ^35^ and all three psilocybin metrics evaluated: PPL, Occ_2A_, and SDI. We observed statistically significant associations at a range of correlation coefficient thresholds that produce graphs with a mean degree from 22 to 38, the previous study reported significant differences at thresholds producing mean degrees from 24 to 35. Characteristic path length is a description of the number of edges that must be traversed to get from any one brain region to another, a putative measure of capacity for information flow. Our results suggest that one of the effects of psilocybin on the brain can be described as a broadening of the histogram of path lengths across region-to-region connections. Notably, this does not imply that the average path length is shorter or longer, only that there is a wider distribution of these across the whole brain. Our convergent results are encouraging considering that the previously reported dataset used a different drug (ayahuasca, which contains MAOIs as well as the psychedelic N,N-dimethyltryptamine) and different imaging parameters, suggesting robustness of the metric. This association with PsiFx was relatively robust to pre-processing strategies, yet was sensitive to GSR, suggesting that acutely increased global connectivity often reported following psychedelic drug administration may directly affect geodesic entropy. We are not aware of other studies evaluating geodesic entropy, so comparison to other drugs or psychiatric conditions are not yet possible and should be evaluated in future studies.

### Dynamic Conditional Correlation Distribution Entropy

We observed a statistically significant positive relation between PsiFx and DCC entropy for all within or between network relations except within the motor network. DCC entropy is a measure of the width of the distribution of instantaneous connectivity values for any region-region edge across each scan. The previous study found no change in DCC distribution at one-week and one-month post administration of psilocybin ^37^ and importantly did not evaluate acute effects. We observed moderate-to-strong correlations with PsiFx (i.e., Pearson’s rho with PPL > 0.7 for three edges, all including DMN (DMN-DMN, DMN-Frontoparietal, DMN-Medial Frontal), and Pearson’s rho > 0.5 for 28/36 network edges). The strength of these associations is remarkable, perhaps as large as any previously reported fMRI effect of psychedelic action, suggesting that DCC distribution may be a strong candidate neural correlate of acute psychedelic effects and among the strongest correlations observed in pharmaco-fMRI. Our results suggest that psilocybin increases the variability of connectivity between regions across time across almost all region-region pairs, which are summarised into networks. Furthermore, this association was robust to most pre-processing strategies. Notably, the association became weakly negative in some network edges when the low-pass filter was removed, suggesting that PsiFx effects on DCC entropy manifest mostly in relatively slow frequencies and are obscured by higher frequencies. As above, we are not aware of other pharmaco-fMRI studies evaluating DCC distribution. Notwithstanding, the sheer magnitude of the observed associations suggests DCC distribution may be a sensitive marker for acute psychedelic effects on the brain and so we encourage independent replication.

### Multi-scale Sample Entropy

We observed a significant positive relation between PsiFx and scale-1 sample entropy (i.e., temporal resolution = 2-seconds) in seven out of 17 brain networks. Conversely, we observed a significant negative relation between scale 5 sample entropy (temporal resolution = 10-seconds) in 14 out of 17 networks. However, these associations at both scales were affected by the denoising pipeline, only maintaining a positive association for two and one additional pipelines respectively and even negative associations at the short scale after removing the low-pass filter. Multi-scale sample entropy measures the irregularity of a signal over its entire length. Increased sample entropy in most networks at scale-1 and decreased sample entropy at scale-5 align in both cases with the original observation following LSD ^29^. fMRI-measured multi-scale sample entropy has been shown to be increased in the default-mode, visual, motor and lateral-prefrontal networks following caffeine ^41^ and decreased at scale 1 during sleep ^38^, although certain parameters used in their calculations were different to those employed here. As such, it is possible that the effects that we, and Lebedev and colleagues, observed may reflect differences in wakefulness and may thus be non-specific to psychedelic effects. Positive symptoms of schizophrenia have been positively associated with sample entropy at scales 1 and 2, and negatively associated in certain brain regions at scales 3, 4, and 5 ^47^. This is consistent with our observations and is also phenomenologically consistent, as the high-dose psychedelic state has some overlap with some positive symptoms of schizophrenia e.g., verisimilitude, alterations in visual perception (though psychedelics do not normally produce ‘true’ hallucinations, i.e., sensory appearances indistinguishable from reality, as are present in schizophrenia), and sense of self. We are aware of the problematic history of psychedelic ‘psychotomimetic’ research and urge caution in overinterpretation of this apparent convergence ^48^. Our convergent results with Lebedev and colleagues are intriguing considering that the original paper reported effects following intravenous LSD administration whereas we administered psilocybin orally. However, the diverging effects of denoising pipelines suggests that this metric is susceptible to retained signal frequencies and global signal. Taken together, increases in sample entropy at short temporal resolutions may be a candidate biomarker for psychedelic effects, if they cannot be explained by, e.g., wakefulness state.

### Lempel-Ziv Complexity

Intriguingly, Lempel-Ziv complexity of two measures (meta-state complexity and temporal BOLD complexity (LZct)) were significantly positively associated across several denoising pipelines. Meta-state complexity was significantly weak-moderately associated with at least one PsiFx in all but one pre-processing pipeline. Furthermore, recent work has shown a negative relation between meta-state complexity and Global Control Energy (GCE), representing the amount of input required to shift from one brain-state to the next ^49^. GCE has been shown to decrease during DMT infusion and associated with LZct of simultaneously collected EEG signals ^50^. This suggests a potential bridge between fMRI meta-state complexity and EEG LZct through the lens of Network Control Theory ^51^. These convergent findings support meta-state complexity as an insightful metric of psychedelic effects on brain activity. However, the original study of meta-state complexity does not report a statistical analysis of intravenous LSD nor intravenous psilocybin effects, but the data are publicly available and do not support a significant effect of either drug ^32^. The original study of BOLD complexity reports an increase in spatial BOLD complexity following LSD, but not psilocybin and does not report any findings pertaining to temporal BOLD complexity ^2^. One MEG and four EEG studies have reported increased LZc following psychedelic administration ^52–56^, providing convergence for its utility as a marker of psychedelic effects, however one reports increases in LZct in the absence of subjective drug effects, indicating a potential epiphenomenon ^56^. Given this intriguing convergence across modalities, future studies should evaluate whether there is a relation between LZct of the EEG and BOLD timeseries. Across previous studies analysing regional timeseries, there is inconsistency in the quantification of LZc in the temporal (LZct) and spatial (LZcs) domain. Our borderline statistically significant associations were observed for LZct only. It is our perspective that LZct is more sensible and should be used in future studies as it preserves region-specific temporal information, whereas LZcs is sensitive to arbitrary region order.

### Null Findings

We consistently did not observe a significant association with PsiFx for 8 of the 14 brain entropy metrics considered here (Table 1). Of these, the original studies reported either increased entropy following psychedelic drug administration ^19,27,28,36^, no effect ^30^, or did not formally evaluate the effect of psychedelic drug administration ^26^ (Table 1, Supplementary Table S1). Our observed entropy estimates for hippocampal-ACC motif entropy are markedly different from those previously reported ^28^. We are concerned that the originally reported values are not mathematically possible, see the Supplementary Text and Supplementary Figure S3 for a detailed consideration. Although our null findings with respect to these metrics does not establish that they have no relation to acute psychedelic drug effects, they imply a smaller relation that limits their utility as biomarkers of acute psychedelic effects. The discrepancy between our observations and those reported previously underscores the need to replicate or corroborate findings in independent cohorts to validate initial reports.

Our inability to replicate previous findings may be due to greater statistical power and different statistical models i.e., linear regression with PsiFx. Incongruence may also be attributed to differences in data collection. All previous studies reporting acute effects on brain entropy following a range of routes of administration and across psychedelic drug classes though the acute effect appear similar, we cannot rule out differences due to drug or route of administration. However, if entropic brain effects are not consistent across drugs this would indicate that these metrics are not useful neural correlates of the psychedelic experience. We encourage all future fMRI studies evaluating psychedelic effects on brain function to measure subjective drug intensity at time of scanning and to collect plasma samples for quantification of plasma drug levels as described in the psychedelic fMRI consensus paper ^16^.

### Inter-correlation Between Entropy Metrics

Despite the large set of brain entropy metrics that have been reported previously, no studies have considered whether these measures are inter-correlated. We observed positive associations between geodesic and degree distribution entropies, which are based on the same graph-theory representation of connectivity, between LZcs and LZct which are conceptually very much related, and between the mathematically similar NGSC and Von Neumann entropy as discussed above. Notably, we observed four pairs of brain entropy metrics that were significantly negatively correlated. This highlights the importance of specificity in describing “brain entropy.” Many of these metrics represent distinctly different constructs, their individual meaning and collective representation of psychedelic effects is muddied by superficially considering them all metrics of “brain entropy.” Future studies should be cognisant of this variable relation in considering whether findings are consistent or convergent across studies.

### Implications for the Entropic Brain Hypothesis

We evaluate this set of metrics in part to address uncertainties around The Entropic Brain Hypothesis, a prominent model that suggests 1) there is a relation between the magnitude of entropy of a brain activity signal and the subjective quality of that conscious state, 2) subjective psychedelic effects stem from a drug-induced increase in brain function signal entropy. (See Figure 7 of Carhart-Harris et al., 2014). Herein we have observed that only certain types of fMRI brain-entropy are associated with psychedelic drug effects. Notably, the EBH does not articulate a specific entropy quantification, our mixed findings highlight a relevant lack of specificity within the hypothesis; it is difficult to clearly say whether we find support for or against it. Our findings do reproduce and identify novel effects, e.g., geodesic entropy, sample entropy, DCC entropy, and NGSC. It is also notable that some metrics advanced as supporting this hypothesis, e.g., LZ complexity, are not measures of entropy, “in the information theoretic sense” as described in the original hypothesis paper, but rather complexity. The limited correlation between measures highlights that the previous studies provide evidence for distinct hypotheses and not necessarily support for a singular model. Extending beyond entropy, future work should consider whether candidate entropy metrics align with other models of psychedelic brain-action such as the REBUS ^70^ and the thalamic gating hypothesis ^71^.

### Alternative Neuroimaging Techniques

Although we focus here on fMRI quantifications of entropy, it is worth noting that psychedelic effects on brain entropy, specifically LZc, have been applied to five original MEG and EEG datasets, over eight papers ^52–59^. The entropy measures applied in these studies leverage the high temporal sampling rate that is not clearly applicable to temporally slower fMRI and were not evaluated here. Further, the methods capture different aspects of physiological response to psychedelics. Future work evaluating psychedelic effects on brain entropy using multimodal neuroimaging and evaluating relations between alternative quantifications of brain-entropy will contribute meaningfully to the field.

### Limitations

Brain imaging data were acquired on two different MRI scanners with different sequences (e.g., different TRs) requiring temporal downsampling of some data to match the other. However, each participant was scanned on only one scanner, enabling us to map within-subject changes onto PsiFx independent of scanner differences. Our study did not include a placebo condition, but we did acquire a pre-drug scan with which we estimated brain entropy metrics in the absence of psilocybin effects. Due to the large within-subject variability in fMRI outcomes in participants scanned several days apart, pre-drug vs post-drug scans performed on the same day may be superior to placebo scans performed weeks apart for evaluating drug effects because it limits this within-subject variance component ^60^. However, we did not administer negative control substances that would allow investigation of specificity. Consequently, we cannot determine which metrics are specific to psychedelics. Future research aiming to identify psychedelic-specific entropy biomarkers should include active comparators from drug classes including stimulants (amphetamine, caffeine), NMDA agonists (ketamine, nitrous oxide), MDMA, and cannabinoids (delta-9-THC). Such comparative studies would clarify whether observed entropy changes reflect serotonergic 5-HT2A agonism specifically or broader alterations in brain state complexity common to various psychoactive substances. PPL and SDI were associated with increased motion in the scanner, see Supplementary Figure S4; although we included an estimate of motion as a covariate in our models, employed scrubbing, motion correction and denoising strategies, and show that motion was not positively associated with any whole-brain entropy metrics, we cannot rule out that motion confounds our reported effects. It has been reported in many groups that head motion is increased following psychedelic drug administration so this is not a limitation unique to our data ^53^. Most fMRI scans were 10 minutes long, though some were only five. This may not be long enough to derive stable estimates of brain entropy metrics, e.g., previous studies have recommended >13 minutes for single-echo fMRI ^61^. Our data include scan sessions of three different volume lengths due to differences in TR and scan duration; notably, all scans for each subject were the same length. Entropy quantifications that preserve the temporal ordering of the time-series, such as sample entropy and the Lempel-Ziv complexity measures may be sensitive to time-series length. Future studies resolving optimal scan parameters for estimating stable brain entropy estimates would benefit this emerging field. Approximately half of the scans analysed herein utilised a multi-band acceleration protocol that may negatively affect signal-to-noise ^62^, though these effects may be less pronounced for task-free imaging as performed here ^63^. We corrected for estimated physiological noise using aCompCor but did not measure physiological effects such as by using eye-tracking or statistically model physiological effects such as changes in respiration, heart rate, or vasoconstriction which are affected by psilocybin and may have confounded our findings ^64,65^. Psilocybin has been shown to increases cerebral blood flow ^66^, and LSD has been shown to decrease it ^67^. Furthermore, the psychedelic DOI has been shown to alter neurovascular coupling i.e., the relation between neural activity and hemodynamic response, in mice ^68^. These haemodynamic effects of psychedelics may alter the BOLD signal in a way that confounds the interpretation of the underlying neural effects. Future research should aim to quantify psychedelic effects on neurovascular coupling in humans and the effect of this on signal complexity. Our statistical models assume a close temporal relation between brain entropy and PsiFx (measured adjacent to scans), thus, if changes in brain entropy occur after PsiFx, they would not be well captured.

### Denoising

A prevailing challenge in fMRI research is how best to handle the enormous flexibility in denoising strategies ^69^. Here we explored this space by evaluating the moderating effect of different denoising strategies on brain entropy metrics. Most of our results were robust to atlas or parcellation choice. The associations with PsiFx of some metrics (DCC entropy, NGSC, meta-state complexity and geodesic entropy) were robust to most denoising strategies, whereas other metrics were sensitive to denoising strategy (e.g., Sample entropy). For those metrics which remained significantly associated with PsiFx across pipelines, the strength of some associations varied across pipelines. Notably, removing the low-pass filter and applying a relatively narrow bandpass filter both substantively affected the statistical relations between PsiFx and brain entropy metrics. This is consistent with previous reports that pre-processing decisions can influence observed effects on fMRI outcome measures^69^, which underscores the need for future studies in large, normative datasets to evaluate brain entropy metric characteristics across this denoising multiverse. This is all the more relevant to advance their predictive or prognostic utility in clinical cohorts. For the purposes of this manuscript, we show that our main denoising pipeline was very similar to all previously applied in this space (See Supplementary Table S7) and thus believe that our results are directly comparable with previously reported findings.

We also present here the Copenhagen Brain Entropy toolbox (CopBET https://github.com/anders-s-olsen/CopBET), a Matlab-based toolbox containing functions to calculate the entropy metrics reported here, supporting the reproducibility of this and future studies. ^69^

## Conclusion

In conclusion, we observed consistent positive effects of psilocybin administration on five considered complexity/entropy metrics: NGSC, DCC entropy, geodesic entropy, meta-state complexity, and sample entropy (scale 1). DCC entropy showed remarkably strong effects, and NGSC demonstrated robust associations across all denoising pipelines. LZct showed inconsistent effects and eight showed very limited or no associations. Limited inter-correlations between metrics and differential denoising sensitivity indicate that these measures capture distinct features of complexity and entropy of BOLD signals, rather than a unitary construct. Our findings provide nuanced support for the Entropic Brain Hypothesis, indicating that not all entropy metrics may reliably capture psychedelic effects. These results establish candidate biomarkers for clinical psychedelic studies and underscore the critical importance of independent replication and methodological transparency in psychedelic neuroimaging research.

## Methods

Twenty-eight healthy volunteers participated in the study (10 female, mean age ± SD : 33 ± 8) and were recruited from a database of individuals interested in participating in a study involving psychedelics. A detailed description of the study design can be found in the Supplementary Text and has been reported previously ^72^. The study protocol was approved by the ethics committee of the capital region of Copenhagen (H-16026898) and the Danish Medicines Agency (EudraCT no.: 2016-004000-61). The study was registered at clincaltrials.gov (NCT03289949). Data presented here were collected between 2018 and 2021. A subset of the functional brain imaging data presented here has been included in different studies reported previously ^72,73^. Details of recruitment, procedures during the psilocybin session, ethical approvals, MRI acquisition and quality control, are described in the Supplementary Text. Analyses were pre-registered on the 3rd of August 2022 (https://aspredicted.org//bw8y7.pdf). Some analyses that met our inclusion criteria (i.e., fMRI studies investigating entropy changes pertaining to psychedelics) were identified after pre-registration and were added. No statistical methods were used to pre-determine sample sizes but our sample sizes are larger than all previous publications (see Figure 1).

### Data Collection

After obtaining written informed consent and screening for neurological, somatic and psychiatric illness, participants completed a single-blind, cross-over study design wherein participants received a single 0.2-0.3 mg/kg dose of psilocybin (mean ± SD dose: 19.7 ± 3.6 mg, administered in units of 3 mg capsules) or 20 mg of ketanserin. Data from ketanserin scans are outside the scope of the current evaluation and not presented here. After drug administration, participants completed MRI scan sessions including resting-state fMRI (see Supplementary Text for details) approximately 40, 80, 130, and 300 minutes after administration. Following each scan, participants were asked, "On a scale from 0 to 10 how intense is your experience right now" to measure SDI and a venous blood-draw used to quantify PPL (see Supplementary Text for details). After each resting-state fMRI scan, participants were asked if they had fallen asleep (no participants reported doing so). Occ_2A_, i.e., occupancy of psilocybin at the 5-HT2A receptor is closely related to PPL and SDI ^2^. Here we applied the previously reported parameter estimates relating PPL to occupancy based on the Hill-Langmuir equation: 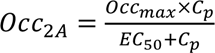 where Occ_max_ refers to the maximum measurable occupancy, C_p_ refers to the measured concentration of the ligand in plasma (i.e., PPL), and EC_50_ refers to the concentration in plasma at which occupancy is equal to 50% of Occ_max_ (fixed parameters used to compute Occ_2A_: EC_50_ = 1.95 µg/L and Occ_max_ = 76.6%).

### Pre-processing

Pre-processing and denoising was uniform across all entropy metrics despite differences in the pipelines of the original publications. Our pipeline included slice-timing correction (where applicable), unwarping, realignment, co-registration of structural scans to functional data, segmentation, normalisation, and smoothing. Two MR-scanners were used to acquire the data, and some functional data were temporally downsampled so that the sampling frequency was consistent across scan sessions. Denoising in CONN ^74^ included linear detrending, aCompCor ^75^, 12-motion (three translations, three rotations and their first derivatives) and artefact-flagged volume regression (z > 4 SDs or motion > 2 mm using ART), band-pass filtering (0.008-0.09 Hz) and parcellation. Cerebellar ROIs were removed from included atlases as they were not consistently within the field of view. See Supplementary Text for more details.

### Entropy Quantifications

All below entropy quantifications are replications of previously reported methods. Where a metric has specific hyperparameters, we ensured that they were applied as in the previous report via communications with the original authors.

### Entropy of Static Connectivity

Four studies evaluated the entropy of static connectivity given by the matrix of Pearson correlation coefficients, **R**, computed from *N*-regional time-series data ^27,34–36^, *N* being the number of ROIs in the atlas used by the study.

#### Out-network Connectivity Distribution Entropy

Following a graph-theory framework, ROIs from the 200-region Craddock-atlas ^76^ were partitioned into “networks” using the Louvain modularity algorithm applied to the average connectivity matrix across scan sessions ^77^. The “Out-network Connectivity", referred to as "diversity coefficient" in the original publication and “Brain Connectivity Toolbox” ^27,78^, of an ROI was calculated for each scan session as the Shannon entropy of the distribution of connectivity estimates between a given ROI and the set of ROIs assigned to a different network.

#### Degree Distribution Entropy

Degree refers to the number of non-zero elements in any given row of a thresholded matrix. ROI-specific degrees are computed based on **R**, the Pearson correlation matrix between ROIs, with *N*=105 using the Harvard-Oxford-105 atlas ^79^. Both this analysis and geodesic distribution entropy use the absolute correlation values. The thresholding for this analysis occurred in two steps. In the first step, any correlation for which the corresponding p-value was above 0.05 was set to 0. In the second step the goal is to reach a pre-specified mean degree across rows. In order to achieve this, a threshold below which all absolute values are set to 0 is gradually increased until the mean number of non-zero elements is at the desired level. Here we applied a scan-specific threshold that produced a mean degree of 27 because this was the threshold that produced the largest effect in the original publication ^36^. This means that each scan may have a different absolute threshold value, but identical mean degree. The final entropy quantification is simply the Shannon entropy of the distribution of degrees across ROIs. We also calculated entropy for mean degrees of 1 up to the point at which for any given scan session an increase in absolute threshold did not produce an increase in mean degree, i.e., 48. This also applies to the geodesic distribution entropy described below.

#### Geodesic Entropy

Again using absolute correlation values, the matrix was thresholded using only the mean-degree criteria and not the p-value threshold. The matrix was then binarised, setting all non-zero elements to 1. The "shortest path length" was then computed as the fewest edges one must traverse to go from one node to another. The Shannon entropy of the distribution of path lengths from each node to all other nodes was then calculated ^35^. Geodesic entropy was evaluated for correlation coefficient thresholds up to a mean degree of 53.

#### Von Neumann Entropy

Entropy of the Pearson correlation matrix, **R**, derived for the Harvard-Oxford-105 atlas, was calculated through the von Neumann entropy: 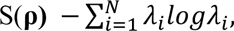 where *λ* are the eigenvalues of the scaled correlation matrix **ρ**=**R**/N. The von Neumann entropy may also be defined as S(**ρ)** = -tr(**ρ**log**ρ)**, where log represents the matrix logarithm ^34^.

#### Normalised Global Spatial Complexity

NGSC was calculated via the eigenvalues of the connectivity matrix computed by a principal component analysis (PCA) of the z-scored time by voxel data matrix. Only voxels within a cortical gray matter mask were used. The eigenvalues were then rescaled by their sum to represent a probability distribution, and the Shannon entropy calculated and rescaled by the maximal value, log(m), where m is the number of non-zero eigenvalues equal to the number of degrees of freedom of the PCA. Notably, disregarding the rescaling by log(m), NGSC is exactly equal to Von Neumann entropy except that the latter used an atlas to reduce the spatial dimensionality of data.

### Entropy of Dynamic Connectivity

#### Intra-network Synchrony Distribution

Nine brain networks were defined according to a previous study ^80^: auditory, dorsal attention, default mode, left and right frontoparietal, motor, salience, visual 1 and visual 2. For a given network, for a given time point, the variance across voxels within the network was evaluated. The Shannon entropy was then calculated on the histogram of the variance estimates over time ^19^.

#### Motif-connectivity Distribution

Dynamic functional brain connectivity was evaluated in four regions (10mm diameter spheres) located at bilateral hippocampi, MNI coordinates: right: (26, -21, -16), left: (-34, -22, -16), and anterior cingulate cortices, right: (4, 35, 18), left: (-2, 23, 28) using a non-overlapping sliding window approach with varying window lengths (15-150s). In each window, the partial correlation coefficient and corresponding p-value was calculated for every region pair, controlling for the remaining regions and the motion framewise displacement time-series. These time-series were standardised before windowing. The 4 x 4 partial correlation matrix was binarised for every window, according to a corrected significance threshold p=0.0083 (i.e., 0.05/6, where 6 is the number of region pairs). A probability distribution of the frequency of each of the 64 possible graph structures was established and the Shannon entropy was calculated ^28^.

#### LEiDA-state Markov-rate

Notably, this entropy metric was not applied to evaluate psychedelic effects in the original paper. Rather, the authors provided a computational framework wherein parameters were learned by optimising this entropy measure. For each scan session, Leading Eigenvector Dynamics Analysis (LEiDA) ^81^ was applied to the time-series of 90 AAL atlas regions ^82^. The phase series was computed using the Hilbert transform and, for each time point, a phase coherence matrix was estimated based on the cosine of the difference between pairwise instantaneous phases. The phase coherence matrices were decomposed using the eigenvalue decomposition and the first eigenvector was retained for every time point. The set of eigenvectors was clustered using *K*-means with *K* = 3 states. Subsequently, the transition probability matrix was computed for each scan session. The entropy rate of the transition matrix, *P*(*i*, *j*), for each state, i, was calculated as 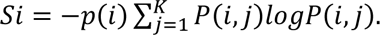.where, *p* is the leading eigenvector of *P*. The final entropy measure is given as 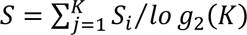 ^26^.

#### Dynamic Conditional Correlation Distribution

Regional time-series were evaluated for each of the regions described in the Shen 268 region atlas ^83^. Windowless framewise correlation coefficients were calculated for all edges using the Dynamic Conditional Correlation (DCC) toolbox ^84^. Subsequently, the probability distribution over each ROI-to-ROI DCC time-series was established, and the Shannon entropy was calculated. Each ROI was assigned to one of eight networks: default mode, fronto-parietal, medial-frontal, motor, subcortical-cerebellar, visual association, visual 1, and visual 2. Each ROI-to-ROI pair was assigned to its respective network-to-network association (e.g., motor-to-motor, default mode-to-motor) and the mean entropy of each network-to-network association was calculated. Although the original publication applies bin-width correction, they do not report an effect of bin width and we report findings using MATLAB’s *histcounts* function, which automatically calculates bin-width ^37^. Thus, we did not implement bin-width correction.

#### Meta-state Complexity

Regional time-series were evaluated for each of the regions described in the Lausanne 463 region atlas ^85^. BOLD time-series across all scan sessions were clustered using *K*-means into *K* = 4 states using the Pearson correlation distance metric. The clustering procedure was repeated 200 times with random initialisations and the best repeat in terms of *K*-means loss was extracted. The four states were grouped into two meta-states because the clustering procedure typically produces sign-symmetric states. Each volume was assigned to meta-state 0 or 1 and the Lempel-Ziv complexity (LZ76 exhaustive algorithm) of this binary sequence was calculated ^32^.

#### Integration/Segregation-state Distribution

Regional time-series were evaluated for each region described in the Schaefer 200 region atlas ^45^, augmented with 32 subcortical regions from the Tian atlas ^44^. A sliding-window correlation analysis was performed using a window defined by convolving a rectangular window of size 44 seconds with a temporal Gaussian kernel (FWHM = 3s). The correlation matrix was established for each window (stride of 1), and the Louvain modularity algorithm ^77^ was applied to estimate the module degree z-score and participation coefficient for each region. The Louvain modularity algorithm was repeated 100 times to ensure an optimal assignment. *K*-means clustering with *K* = 2 states was applied to a cartographic profile, i.e., a two-dimensional unnormalised histogram of these measures, using the correlation distance and 500 replications. The Shannon entropy was computed on the probability distribution of state occurrences ^30^.

### Entropy of Regional Dynamics

#### Multi-scale Sample Entropy

Networks were defined using the Yeo 17-network atlas ^86^. Sample entropy is defined as the negative logarithm of the conditional probability that if two vectors with length *m* (set to 2) are dissimilar below a threshold distance *r* (set as 0.3), then vector pairs with length *m* + 1 will also have distance below the threshold ^21^. Scales 1-5 were evaluated for each network, meaning that each time-series was split into non-overlapping windows of length (scale) *s* volumes and the means of each window were concatenated to form a condensed time-series upon which sample entropy was calculated ^29^.

#### BOLD Complexity

Regional time-series were evaluated for each of the regions described in the Schaefer 1000 region atlas ^45^. BOLD time-series for each ROI were first Hilbert-transformed. The amplitude of the Hilbert series was then binarised around the mean amplitude for that region, i.e., assigned as “1” if greater than the mean and “0” if less. These binarised time-series were combined into an *T* × *N* matrix, where *N* = 1000 is the number of regions and *T* and is the number of time points.This matrix was collapsed into a single vector to compute 1) the Lempel-Ziv complexity over time (LZct, LZ78 algorithm) wherein regional time-series were concatenated or 2) Lempel-Ziv complexity over space (LZcs) wherein time-adjacent “region series” were concatenated. LZct represents a calculation of the temporal entropy of each ROI, whereas LZcs represents a calculation of the spatial entropy at each timepoint. The original publication ^33^ reported only LZcs, but LZct is also described in ^52^ whom Varley and colleagues reference as the source of their methods.

### Statistical Model

Effects of psilocybin on brain entropy metrics were estimated using a linear mixed effects model with relevant R packages, i.e., *predictmeans* (v1.0.6), *lme4* (v1.1.30), *nlme* (v3.1.157), *lmerTest* (v3.1.3) and *LMMstar* (v0.7.6). We regressed each metric against each of the three measures (PPL, SDI, or Occ_2A_) separately with a subject-specific random intercept and adjusting for motion, age, sex, and scanner. A test statistic for the association between metric and measure was obtained using the Wald statistic. To ensure adequate control of the family-wise error rate (FWER) across regions within each of the 14 metrics, (e.g., 17 networks for one time scale of multi-scale sample entropy), we calculate p_FWER_ adjusted using the maxT test method ^43^ in a permutation framework similar to ^87^, employing 10000 permutations. As such, if observed data superseded all permutations, the p-value is reported as p < 0.0001. “Motion” reflects the framewise displacement computed using the Artifact Detection Toolbox (ART) (see Supplementary Text) and “scanner” controls for MR scanner, of which there were two. We do not adjust p-values across metrics, nor across SDI, PPL and Occ_2A_; unadjusted p-values are reported for non-regional metrics as p_perm_. We defined findings as statistically significant if they were associated with all three psilocybin effects, SDI, PPL and Occ_2A_ (collectively summarised “PsiFx”) at p_perm_ < 0.05 for non-regional metrics or p_FWER_ < 0.05 for regional metrics. Effect sizes are reported as Pearson’s correlation coefficient between the partial residuals of the entropy metrics (adjusted for covariates using the mixed-model described above) and each of PsiFx. The strength of Pearson’s correlation coefficients for significant associations are described as “weak” (≤0.3), “moderate” (>0.3 and ≤0.6), or “strong” (>0.6) as previously defined ^88^.

### Moderating Effect of Scanner

Our data were collected on one of two MRI scanners. In order to investigate whether scanner choice had an impact on the estimated relation between PPL and entropy moderating effects of scanner were explored in separate models that included the scanner-x-PPL interaction as an additional covariate, i.e., entropy ∼ PPL + FD + Age + Sex + scanner + PPL:scanner.

### Effect of Parcellation

To explore parcellation effects on outcomes, all entropy metrics were evaluated using the Schaefer 100 region atlas with 16 subcortical regions from the Tian atlas ^44,45^. For metrics using network definitions, the Yeo 7-network atlas was applied as a common atlas.

### Correlation Between Metrics

Simple Pearson correlation coefficients were calculated between each whole-brain entropy metric pair using the common atlas described above. Of the two graph theory metrics requiring thresholding, the threshold producing a mean degree of 27 was used. For the motif-connectivity distribution, 15 and 100 second windows were selected to represent fast and slow dynamics, respectively. P-values were adjusted using Bonferroni correction ^89^. All scans remaining after pre-processing were used in these analyses.

### Effect of Denoising Pipelines

To explore the effect of denoising decisions on the associations between PsiFx and brain entropy metrics, analyses were repeated for six additional pre-processing pipelines. Each pre-processing pipeline was run on the data parcellated as described in the section “Effect of parcellation” i.e., 116 ROIs assigned to seven networks. Each pipeline changed one variable from the main pipeline. These were as follows: 1) adding global signal regression, 2) removing the low-pass 0.09 Hz filter (i.e., not removing high-frequency signal), 3) expanding the 12-motion regressors to include squares of the derivatives (i.e., Volterra expansion), 4) not regressing out flagged volumes 5) regressing flagged volumes with a stricter threshold (z >3 or motion>0.5mm), (6) applying a narrower bandpass filter (0.03-0.07 Hz).

## Code and Data Availability

We shared relevant analysis scripts with original authors, hoping to ensure as much as possible that our computations aligned with original reports; we are thankful for the feedback we received. All functions used to derive entropy estimates from pre-processed data have been compiled into the "Copenhagen Brain Entropy Toolbox" (CopBET), a Matlab-based toolbox that can be found here: https://github.com/anders-s-olsen/CopBET. The permutation testing code is also available here. Code for other statistical analyses and figures can be made available upon request. The data that support the findings of this study are available from the corresponding author upon request to the CIMBI database ^90^.

## Acknowledgements

A sincere thank you to Andrea Luppi, Enzo Tagliazucchi, Alexander Lebedev, Manoj Doss, Thomas Varley, and especially Parker Singleton for their advice and feedback regarding their entropy metrics, and ongoing feedback on this project. We appreciate the advice from Drs Doss and Varley to not use their entropy metrics, as they no longer believed them to be valid, which we appreciate, but disregarded.

## Conflict Statement

DEWM salary was supported by an unrestricted grant from COMPASS Pathways who had no involvement in the preparation or conception of this manuscript or related data collection. MKM has received an honorarium as a speaker for H. Lundbeck. GMK has served as a consultant for Sanos, Gilgamesh, Onsero, Pangea, Abbvie, PureHealthTech, and has received honoraria as speaker for H. Lundbeck and Sage Therapeutics.

## Author Contributions

DEWM conceptualised the manuscript idea, collected data, performed analyses, and wrote the manuscript. ASO conceptualised the manuscript idea, performed analyses, prepared the CopBET, and wrote the manuscript. BO supported statistical analyses and related software. DSS facilitated, supervised, and performed data collection. SA performed data collection. MKM conceptualised the original study, and facilitated and performed data collection. GMK obtained core study funding, conceptualised the original study, and provided feedback on the project. PMF conceptualised the original study, facilitated data collection, and supervised data analysis and manuscript writing. All co-authors reviewed the manuscript, provided feedback, and approved the final version.

## Notes

### Competing Interest Statement

DEWM salary was supported by an unrestricted grant from COMPASS Pathways who have no involvement in the preparation or conception of this manuscript or related data collection. MKM has received an honorarium as a speaker for H. Lundbeck. GMK has served as a consultant for Sanos, Gilgamesh, Onsero, Pangea, Abbvie, PureHealthTech, and has received honoraria as speaker for H. Lundbeck and Sage Therapeutics

### Clinical Trial

NCT03289949

### Clinical Protocols

https://aspredicted.org/bw8y7.pdf

### Funding Statement

The work was supported by Innovation Fund Denmark (grant number 4108 –00004B), Independent Research Fund Denmark (grant number 6110 –00518B), and Ester M. og Konrad Kristian Sigurdssons Dyrevaernsfond (grant number 850–22–55,166–17–LNG). MKM was supported through a stipend from Rigshospitalets Research Council (grant number R130–A5324). BO has received funding from the European Unions Horizon 2020 research and innovation program under the Marie Sklodowska –Curie grant agreement No 746,850. DEM salary is supported by COMPASS Pathways Ltd. Funding agencies did not impact the study and played no role in manuscript preparation and submission.

### Author Declarations

The study protocol was approved by the ethics committee of the capital region of Copenhagen (H-16026898) and the Danish Medicines Agency (EudraCT no.: 2016-004000-61). The study was registered at clincaltrials.gov (NCT03289949).

### Summary of Updates

Added NGSC quantification and reworked the discussion

